# Automated Detection of Dental Caries and Bone Loss on Periapical and Bitewing Radiographs using a YOLO Based Deep Learning Model

**DOI:** 10.64898/2026.04.12.26350726

**Authors:** Hend Alqaderi, Utsavi Kapadia, Yash Brahmbhatt, Aikaterini Papathanasiou, Dara Rodgers, Peter Arsenault, Justin Cardarelli, Athanasios Zavras, Hengxu Li

## Abstract

**Background:** Dental caries and periodontal disease represent the most prevalent global oral health conditions, collectively affecting several billion people. The diagnostic interpretation of dental radiographs, a cornerstone of modern dentistry, is associated with considerable inter-observer variability. In routine clinical practice, clinicians are required to evaluate a high volume of radiographic images daily, a cognitively demanding task in which diagnostic fatigue, time constraints, and the inherent complexity of overlapping anatomical structures can lead to the inadvertent oversight of early-stage pathologies. Artificial intelligence (AI) offers a transformative opportunity to augment clinical decision-making by providing rapid, objective, and consistent radiographic analysis, thereby serving as a tireless adjunct capable of flagging findings that may be missed during routine human inspection.

**Methods:** This study developed and validated a deep learning system for the automated detection of dental caries and alveolar bone loss using a dataset of 1,063 periapical and bitewing radiographs. Two separate YOLOv8s object detection models were trained and evaluated using a rigorous 5-fold cross-validation methodology. To align with the clinical use-case of a screening tool where high sensitivity is paramount, a custom image-level evaluation criterion was employed: a true positive was recorded if any predicted bounding box had a Jaccard Index (IoU) > 0 with any ground truth annotation. Model performance was systematically evaluated at confidence thresholds of 0.10 and 0.05.

**Results:** At a confidence threshold of 0.05, the caries detection model achieved a mean precision of 84.41% (±0.72%), recall of 85.97% (±4.72%), and an F1-score of 85.13% (±2.61%). The alveolar bone loss model demonstrated exceptionally high performance, with a mean precision of 95.47% (±0.94%), recall of 98.60% (±0.49%), and an F1-score of 97.00% (±0.46%).

**Conclusion:** The YOLOv8-based models demonstrated high accuracy and high sensitivity for detecting dental caries and alveolar bone loss on periapical radiographs. The system shows significant potential as a reliable automated assistant for dental practitioners, helping to improve diagnostic consistency, reduce the risk of missed pathology, and ultimately enhance the standard of patient care.

## 1. Introduction

Oral diseases remain a pervasive global health issue. According to the World Health Organization (WHO), untreated dental caries in permanent teeth is the single most common health condition, affecting an estimated 2.5 billion people, while severe periodontal disease, a primary cause of alveolar bone loss and subsequent tooth loss, impacts approximately 1 billion people worldwide [1,2]. Given that oral disease is affecting systemic health, the high prevalence of these conditions places a substantial burden on healthcare systems and significantly impacts quality of life, underscoring the critical need for timely and accurate diagnosis to enable early intervention and prevent disease progression [3].

Dental radiography is a cornerstone of modern dental practice, providing essential diagnostic information on hard tissues such as teeth and bone that may not be visible during a clinical examination. Periapical and bitewing radiographs are routinely used to detect dental caries and assess alveolar bone levels [4]. However, the diagnostic process is not always without its challenges. In high-volume clinical settings, dentists face significant time constraints and cognitive demands. This pressure, combined with the inherent subtlety of early-stage pathologies on radiographs, may lead to diagnostic oversights. While clinicians are often focused on addressing a patient’s primary complaint, other incipient conditions may be inadvertently missed, delaying crucial treatment.

Interpretation of dental images is a complex perceptual task that relies heavily on the clinician’s experience, training, and visual acuity. This subjectivity can lead to significant inter-observer and intra-observer variability, with studies showing diagnostic discrepancies among dentists for the same radiograph [5,6]. Such variability can result in both missed diagnoses, delaying necessary treatment, and over-treatment, leading to unnecessary procedures.

In recent years, the field of artificial intelligence (AI), particularly deep learning using convolutional neural networks (CNNs), has shown remarkable success in medical image analysis, offering the potential to mitigate these challenges [7]. These technologies can serve as a robust ‘second reader,’ augmenting the capabilities of human experts by providing objective, reproducible, and instantaneous analysis. For dental applications, a growing body of literature has explored the use of CNNs for detecting caries [8,9] and periodontal bone loss [10,11]. The You Only Look Once (YOLO) family of object detection models has become particularly prominent due to its state-of-the-art balance of speed and accuracy, with recent versions like YOLOv8 being successfully applied to various dental diagnostic tasks [12,13]. Crucially, as the proliferation of dental AI research accelerates, there is an urgent need to move beyond isolated performance metrics and ensure models are developed according to rigorous, standardized quality assurance frameworks. To distinguish this work from the broader literature and ensure the highest level of methodological integrity, this study explicitly incorporates and validates its methodology against the newly established ANSI/ADA Standard No. 1110-1 (2025) for Dentistry; Validation Dataset Guidance for Image Analysis Systems Using Artificial Intelligence [16]. By systematically cross-referencing our data collection, annotation, and evaluation protocols against the ADA’s Quality Assurance checklist, this research provides a standardized, reproducible template for clinical AI development.

This study aims to develop and validate an AI deep learning system using the YOLOv8s architecture for the automated detection of both dental caries and alveolar bone loss on periapical radiographs.

In parallel with the technical objectives, this work also served as a vital capacity-building initiative within our dental school. As the integration of AI into healthcare becomes inevitable, it is imperative that the next generation of dental professionals possesses a foundational understanding of these technologies [17]. While annotations were initiated by a group of dental faculty members, the project evolved to provide a hands-on learning experience for dental students, who were actively involved in the data collection, annotation, and validation processes. By embedding AI research within the dental curriculum, we aim to foster AI literacy and equip future clinicians with the skills to critically appraise, validate, and effectively utilize these emerging tools in their practice. This approach aligns with recent calls to establish core AI education curricula within dental medicine to ensure the responsible and effective integration of AI into the profession [18,19].

## 2. Materials and Methods

### 2.1. Ethical Considerations

The study adheres to the ethical standards outlined by the World Medical Association (Declaration of Helsinki) and strictly follows the ANSI/ADA Standard No. 1110-1 (2025) for Dentistry-Validation Dataset Guidance for Image Analysis Systems Using Artificial Intelligence [16]. Ethical approval for the study was obtained from Tufts University’s Institutional Review Board (IRB), under protocol # MOD-07-STUDY00004957, which addressed the de-identification of data, data storage, HIPAA compliance, and data transmission protocols. In alignment with the ADA standard’s AI governance and ethics framework, this study acknowledges the critical importance of mitigating automation bias and ensuring human oversight. The AI system is designed as an assistive clinical decision support tool, not an autonomous diagnostic agent, thereby maintaining clinician accountability and minimizing the risk of patient harm from potential false negatives or false positives.

### 2.2. Data Collection and Preparation

#### Data Sources and Eligibility Criteria

The overall methodology of this study is depicted in the workflow diagram in Figure 1.

**Figure 1.**
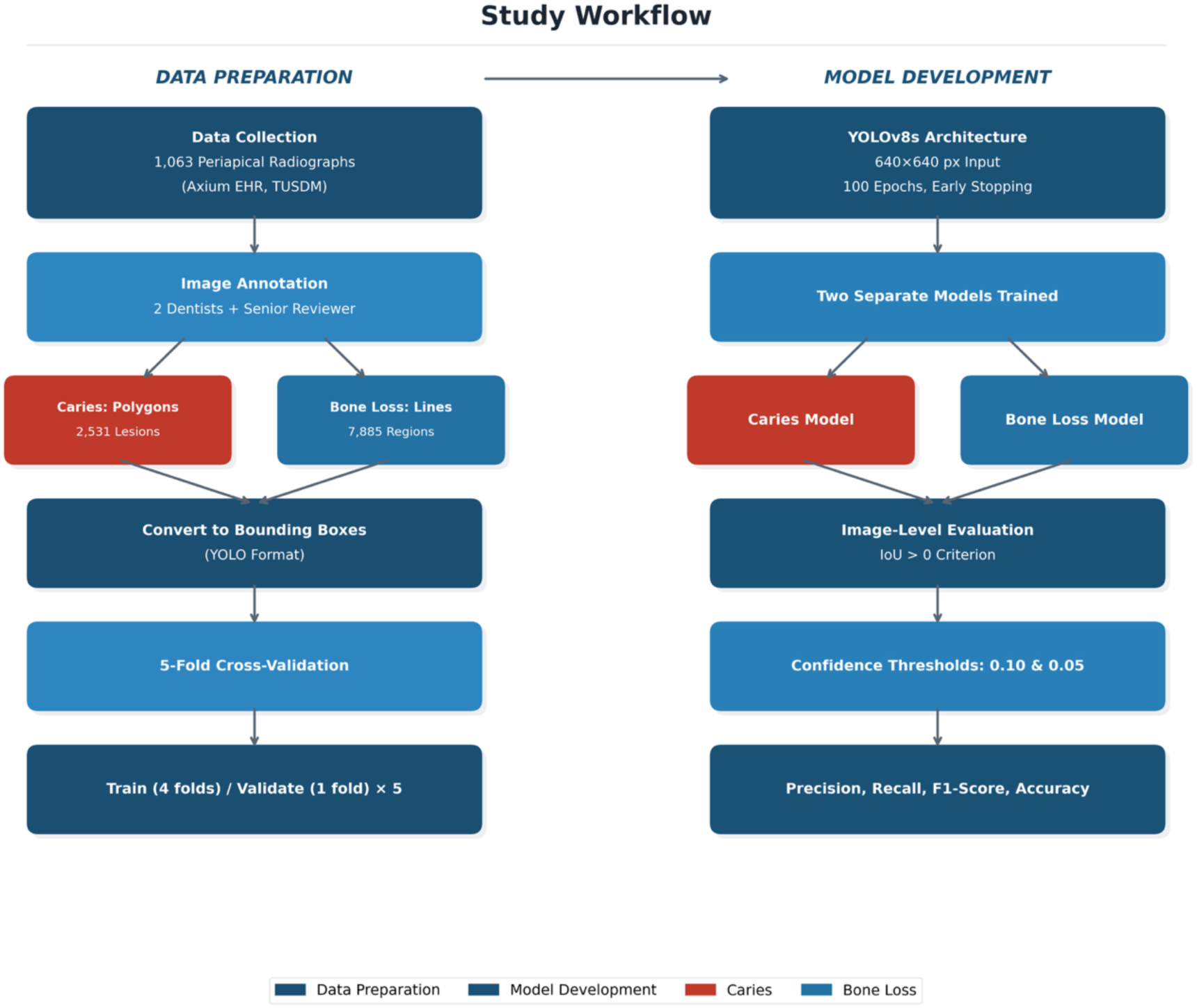
Flowchart illustrating the complete study workflow, from data collection and annotation on the left, to model development, training, and evaluation on the right.

A retrospective data collection was performed using the Axium Electronic Health Records (EHR) database at Tufts University School of Dental Medicine (TUSDM). Patient records from January 2020 to May 2024 were queried. The inclusion criteria were as follows:

- Patients aged 19 years or older.
- Charts containing procedural codes for amalgam or composite restorations (D2140, D2150, D2160, D2391, D2392, D2393, D2394).
- Availability of one or more periapical or bitewing radiographs associated with the restoration codes.

#### Data Extraction and Preprocessing

Four dental students manually extracted the required data, which included patient demographics, tooth number, and date of the radiograph. Screenshots of the radiographs were captured from the Axium EHR. While the ANSI/ADA Standard No. 1110-1 recommends the use of native DICOM files in their least processed state to ensure referential integrity, the retrospective nature of this EHR-based study necessitated the use of high-quality screenshots. To align with the image quality principles of ANSI/ADA Standards 1094 and 1099, radiographs with significant image distortion, poor contrast, or where tooth overlap obscured a clear view of the coronal part of the tooth were rigorously excluded from the final dataset. During model development, input images were resized to 640 × 640 pixels for training and inference.

### 2.3. Image Annotation

The final dataset for this study consisted of 1,063 de-identified periapical and bitewing radiographs.

The annotation process was conducted in two stages. First, a group of faculties developed annotation criteria and annotated 100 images individually and 100 images collectively.

Lessons learned were adapted and annotations proceeded by two experienced dental practitioners. Superannuate application was utilized for the annotation process and was conducted by the two dental practitioners using a rigorous cross-over methodology to ensure high data labeling quality. First, both dentists independently annotated the entire dataset.

Subsequently, the datasets were exchanged. The first dentist reviewed the second dentist’s annotations, and vice versa. Following this cross-over review, both practitioners jointly evaluated the dataset again to double-check all labels. Any disagreements were discussed until a final consensus was reached for every annotation. To ensure high data labeling quality and consistency with the ANSI/ADA Standard No. 1110-1, the annotation methodology mapped directly to the standard’s approved techniques (Table 1). For dental caries, annotators utilized Polygon Annotation (Method #9) to delineate precise boundaries around the lesions. For alveolar bone loss, they employed Polyline Annotation (Method #10) to mark the extent of the bone loss from the cementoenamel junction (CEJ). These annotations were subsequently converted to 2D Bounding Boxes (Method #6) for YOLOv8 training.

**Table 1.** Compliance cross-reference of the study’s methodology against the ANSI/ADA Standard No. 1110-1 (2025) Quality Assurance criteria.

| **ADA QA Domain** | **ADA Requirement** | **Compliance Status in Study** | **Section** |
| --- | --- | --- | --- |
| *0. AI Governance* | Ethical framework, risk management, data quality process | COMPLIANT: IRB approval, ADA ethics framework referenced; risks discussed; system designed as assistive tool. | 2.1, 2.2, 4 |
| *1. Image Annotation Errors* | Image quality (ANSI/ADA 1094, 1099), handling occlusions and overlapping annotations | COMPLIANT: Distorted/overlapping images excluded; consensus review for overlapping annotations; methods mapped to ADA Table 1. | 2.2, 2.3 |
| *2. Image Annotation Machine Translation* | Standardized terminology, AI hallucination detection | PARTIALLY COMPLIANT: ADA-defined terms used; false positive analysis performed, but explicit hallucination analysis recommended for future work. | 2.6, 3 |
| *3. Image Annotation Training* | Duplicate image checking | COMPLIANT: Unique patient records queried; images extracted from individual charts. | 2.2 |
| *4. Data Collection Quality* | Uniformity, consistency (IAA), comprehensiveness, relevancy, bias mitigation | PARTIALLY COMPLIANT: Single-institution dataset ensures uniformity; clinical relevance defined. Limitation noted: formal IAA metrics not calculated (rigorous cross-over consensus used). | 2.2, 2.3, 2.5, 4 |
| *5. Entity Annotation* | Clinical context of annotations | COMPLIANT: Tooth-level annotations with clinical context (caries lesions, bone loss from CEJ). | 2.3 |
| *6. Image Data Labeling Quality* | Minimum F1 Score ≥ 0.70 | EXCEEDS REQUIREMENT: Caries F1 = 0.85; Bone Loss F1 = 0.97. | 3 |

Overlapping annotations were resolved through consensus review. This process resulted in a final dataset containing 877 images with 2,531 annotated caries lesions and 1,001 images with 7,885 annotated bone loss regions. During annotation, brightness and contrast adjustments were allowed to improve visualization.

### 2.4. Model Architecture

This study utilized the YOLOv8s model, a state-of-the-art, real-time object detection model developed by Ultralytics [14]. The selection of this architecture aligns with the ANSI/ADA Standard No. 1110-1, which endorses Convolutional Neural Networks (CNNs) as an evidenced-based reference architecture for dental image analysis. Furthermore, the YOLO (You Only Look Once) framework is explicitly recognized in the standard (Table 1, Method #6) as an approved Lifelong Learning (LLL) methodology. YOLOv8 represents an evolution in the YOLO series, incorporating an anchor-free detection head and a C2f module that replaces the C3 module from YOLOv5. These enhancements provide a more efficient feature gradient flow and contribute to the model’s superior balance of high accuracy and computational efficiency. Given the distinct visual characteristics of caries and bone loss, two separate YOLOv8s models were trained independently: one dedicated to caries detection and one to bone loss detection.

### 2.5. Experimental Setup and Training

A robust 5-fold cross-validation strategy was implemented to train and evaluate the models, following established best practices for medical imaging AI development [15]. The dataset was partitioned into five balanced, stratified folds to ensure that each fold contained a representative distribution of images with and without pathology. For each of the five runs, one-fold was held out for validation while the remaining four were used for training. This process was repeated until every fold had served as the validation set once.

Models were trained for a maximum of 100 epochs using a batch size of 16. Early stopping with a patience of 20 epochs was used to prevent overfitting. Training was implemented in Ultralytics YOLOv8 with automatic optimizer selection, which resulted in an AdamW optimizer with an initial learning rate of 0.002 and momentum of 0.9. Training was conducted on a high-performance computing environment equipped with an NVIDIA A100 GPU. Fixed random seeds were used to support reproducibility.

### 2.6. Evaluation Metrics

Performance was assessed at the image level, reflecting a clinician’s evaluation of the radiographs in determining whether pathology is present in a given radiograph. For this study, a custom evaluation criterion was defined to prioritize sensitivity, which is paramount for a screening tool designed to minimize missed diagnoses.

- True Positive (TP): An image containing at least one ground truth label that was correctly identified by the model. A correct identification required at least one predicted bounding box to have a Jaccard Index (Intersection over Union, IoU) > 0 with any ground truth bounding box (derived from the original polygon and line annotations). This lenient criterion ensures that any plausible detection is counted, aligning with a screening-first approach.
- False Positive (FP): An image with no ground truth labels that the model incorrectly predicted as having pathology.
- False Negative (FN): An image containing at least one ground truth label that the model failed to detect entirely.
- True Negative (TN): An image with no ground truth labels that the model correctly identified as having no pathology.

Based on these image-level counts, we calculated Precision, Recall (Sensitivity), F1-Score, and Accuracy. Results were analyzed at two different confidence thresholds (0.10 and 0.05) to observe the trade-off between precision and recall.

## 3. Results

### 3.1. Model Training and Cross-Validation Stability

The YOLOv8s models for both dental caries and alveolar bone loss were trained and evaluated across all 10 independent runs (5 folds for each of the two models). Training convergence was consistently achieved within the 100-epoch limit, with early stopping triggered appropriately to prevent overfitting. The performance of both models was found to be highly stable and robust across the diverse data partitions. This stability is evidenced by the low standard deviations and coefficients of variation (CV) observed across the 5-fold cross-validation. At the primary screening threshold of 0.05, the caries model exhibited a recall CV of 5.49%, while the bone loss model demonstrated an exceptionally tight recall CV of 0.50%. The detailed per-fold performance metrics, illustrating this consistency across all validation folds, are visualized in Figure 4.

### 3.2. Primary Screening Performance (Confidence Threshold = 0.05)

To align with the clinical objective of a highly sensitive screening tool designed to minimize missed pathologies, the primary evaluation was conducted at a confidence threshold of 0.05. The aggregated image-level performance metrics are presented in Table 2 and Figure 2.

**Figure 2.**
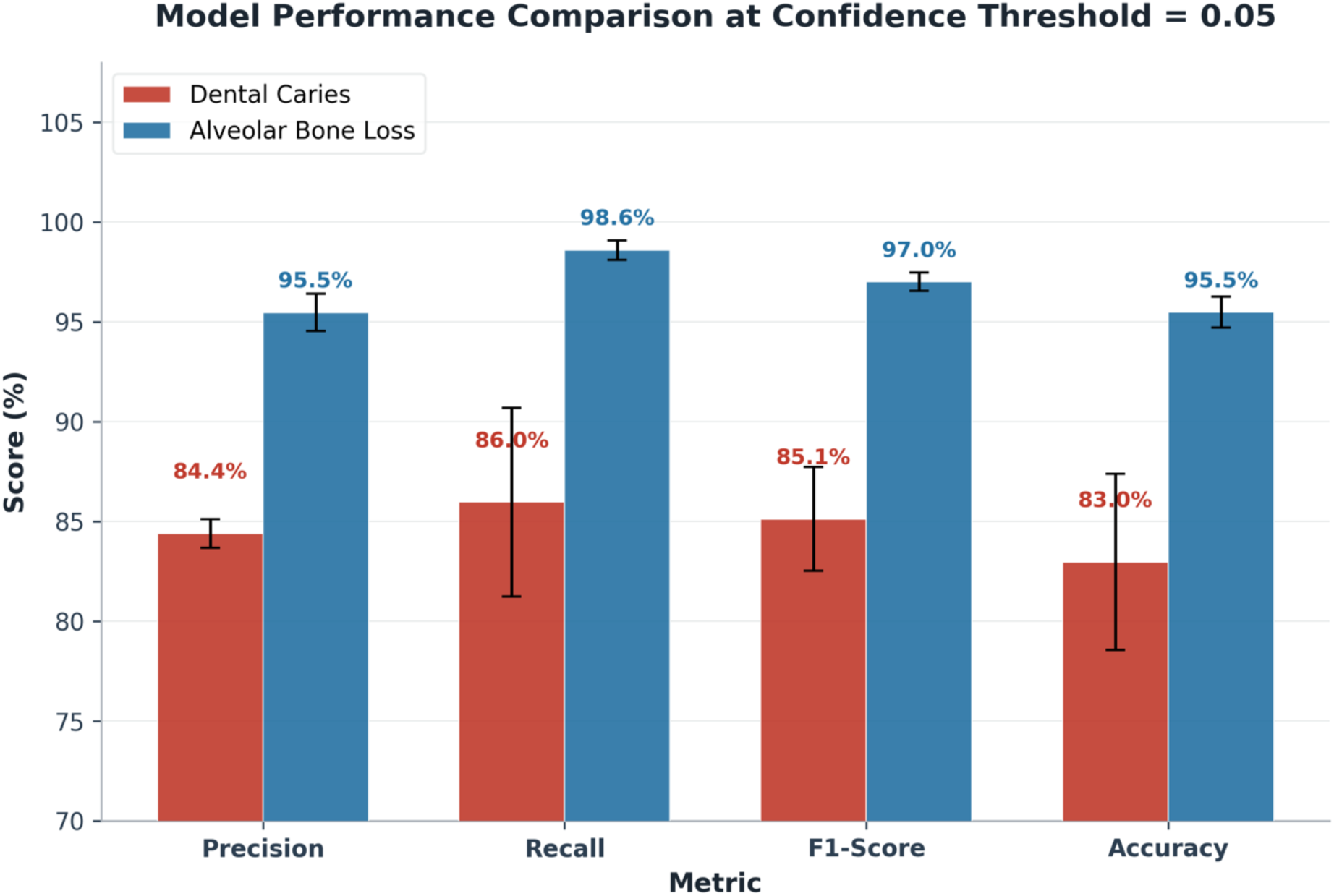
A comparison of the primary performance metrics for the Dental Caries (red) and Alveolar Bone Loss (blue) models at the clinically-oriented confidence threshold of 0.05. Error bars represent the standard deviation across the 5 validation folds.

**Table 2:** Mean image-level performance metrics and standard deviation (±SD) across 5 folds for caries and bone loss detection models at confidence thresholds of 0.10 and 0.05.

| MODEL | CONFIDENCE | PRECISION (%) | RECALL (%) | F1-SCORE (%) | ACCURACY (%) |
| --- | --- | --- | --- | --- | --- |
| CARIES | 0.10 | 90.17 ± 3.41 | 79.98 ± 4.78 | 84.75 ± 3.29 | 83.06 ± 3.42 |
|  | **0.05** | 84.41 ± 0.72 | **85.97 ± 4.72** | 85.13 ± 2.61 | **82.97 ± 4.41** |
| BONE LOSS | 0.10 | 96.17 ± 0.77 | 98.10 ± 0.60 | 97.12 ± 0.45 | 95.76 ± 0.70 |
|  | **0.05** | 95.47 ± 0.94 | **98.60 ± 0.49** | 97.00 ± 0.46 | **95.48 ± 0.78** |

Under these parameters, the alveolar bone loss model demonstrated near-perfect detection capabilities, achieving a mean recall (sensitivity) of 98.60% (±0.49%) and a mean precision of 95.47% (±0.94%). This resulted in an overall F1-score of 97.00% (±0.46%) and an accuracy of 95.48% (±0.78%). The aggregated confusion matrix (Figure 3B) reveals that out of the 1,063 images evaluated across all folds, the model correctly identified bone loss in the vast majority of cases, with an exceedingly low false-negative rate.

**Figure 3:**
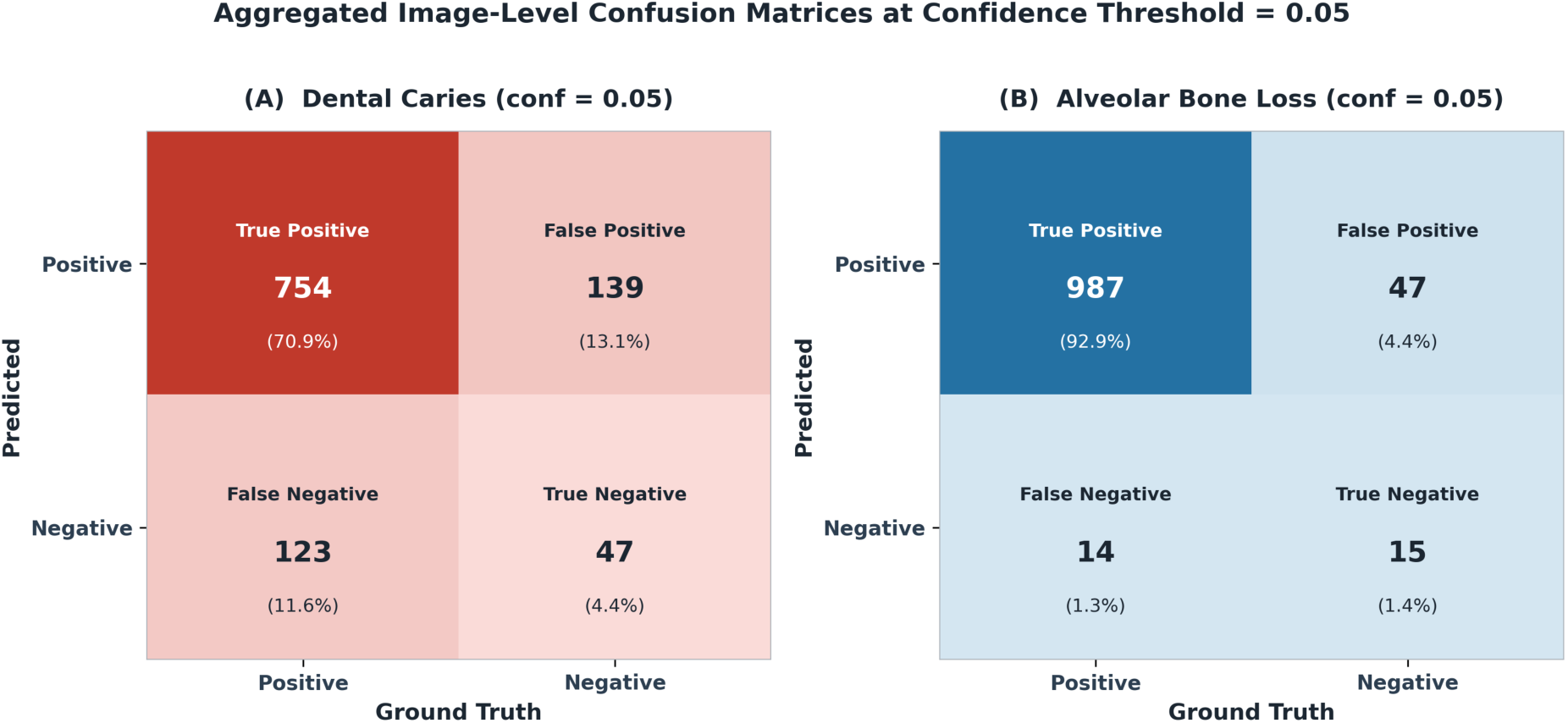
Aggregated image-level confusion matrices at conf=0.05: Aggregated confusion matrices for the caries (Aand bone loss (B) models at a confidence threshold of 0.05. Values are summed across all 5 validation folds. Percentages represent the proportion of the total 1,063 images.

**Figure 4:**
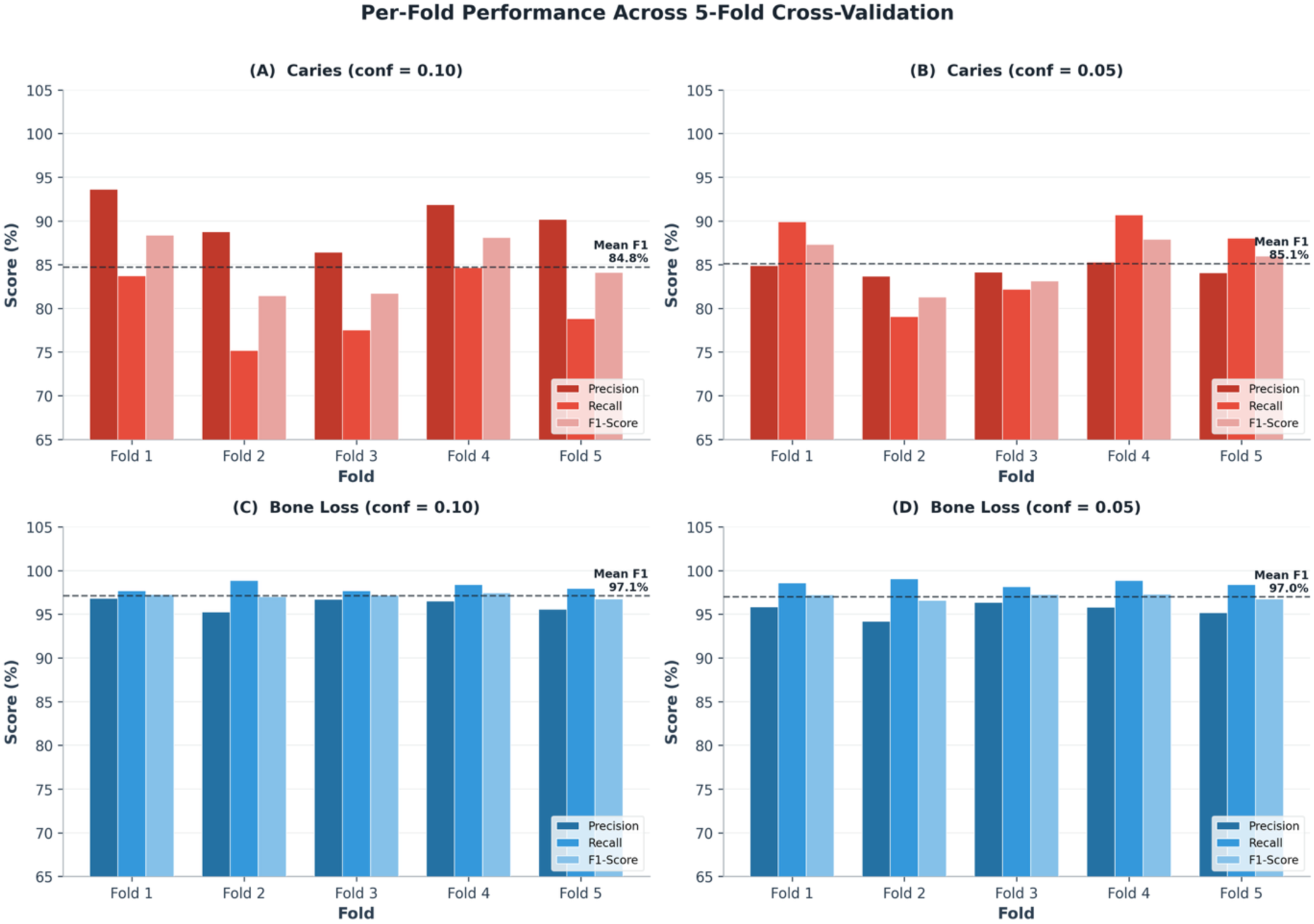
Per-fold performance across 5-fold cross-validation: Per-fold performance across 5-fold cross-validation for both models at both confidence thresholds. The bar charts illustrate the consistency of precision, recall, and F1-score across all folds. The dashed line indicates the mean F1-score.

The dental caries model also exhibited strong performance, achieving a mean recall of 85.97% (±4.72%) and a mean precision of 84.41% (±0.72%). The resulting F1-score of 85.13% (±2.61%) and accuracy of 82.97% (±4.41%) indicate an effective detection system, particularly given the subtle and morphologically variable radiographic presentation of early-stage carious lesions. The aggregated confusion matrix for caries (Figure 3A) demonstrates the model’s ability to correctly flag positive cases while maintaining a manageable false-positive rate.

Crucially, the F1-scores for both the caries (85.13%) and bone loss (97.00%) models significantly exceed the minimum performance threshold of 0.70 mandated by the ANSI/ADA Standard No. 1110-1 Quality Assurance criteria (Table 1, Item 6a), validating the models’ high data labeling quality and predictive reliability.

### 3.3. Precision–Recall Trade-off (Confidence Threshold = 0.10)

To comprehensively evaluate the models’ behavior and illustrate the inherent clinical trade-off between sensitivity and specificity, performance was also analyzed at a stricter confidence threshold of 0.10 (Table 2). As anticipated, increasing the confidence threshold resulted in higher precision at the expense of recall. For the caries model, precision increased to 90.17% (±3.41%), while recall decreased to 79.98% (±4.78%). Similarly, the bone loss model saw precision rise to 96.17% (±0.77%), with a slight reduction in recall to 98.10% (±0.60%). The full Precision–Recall curves (Figure 5) further demonstrate that both models—particularly the bone loss detector—maintain high precision even as recall increases, indicating robust discriminative power across a range of operating points. The selection of the 0.05 threshold for the primary results reflects a deliberate optimization for sensitivity, ensuring the AI system functions effectively as a ‘second reader’ that prioritizes the flagging of all potential lesions for subsequent clinician review.

**Figure 5:**
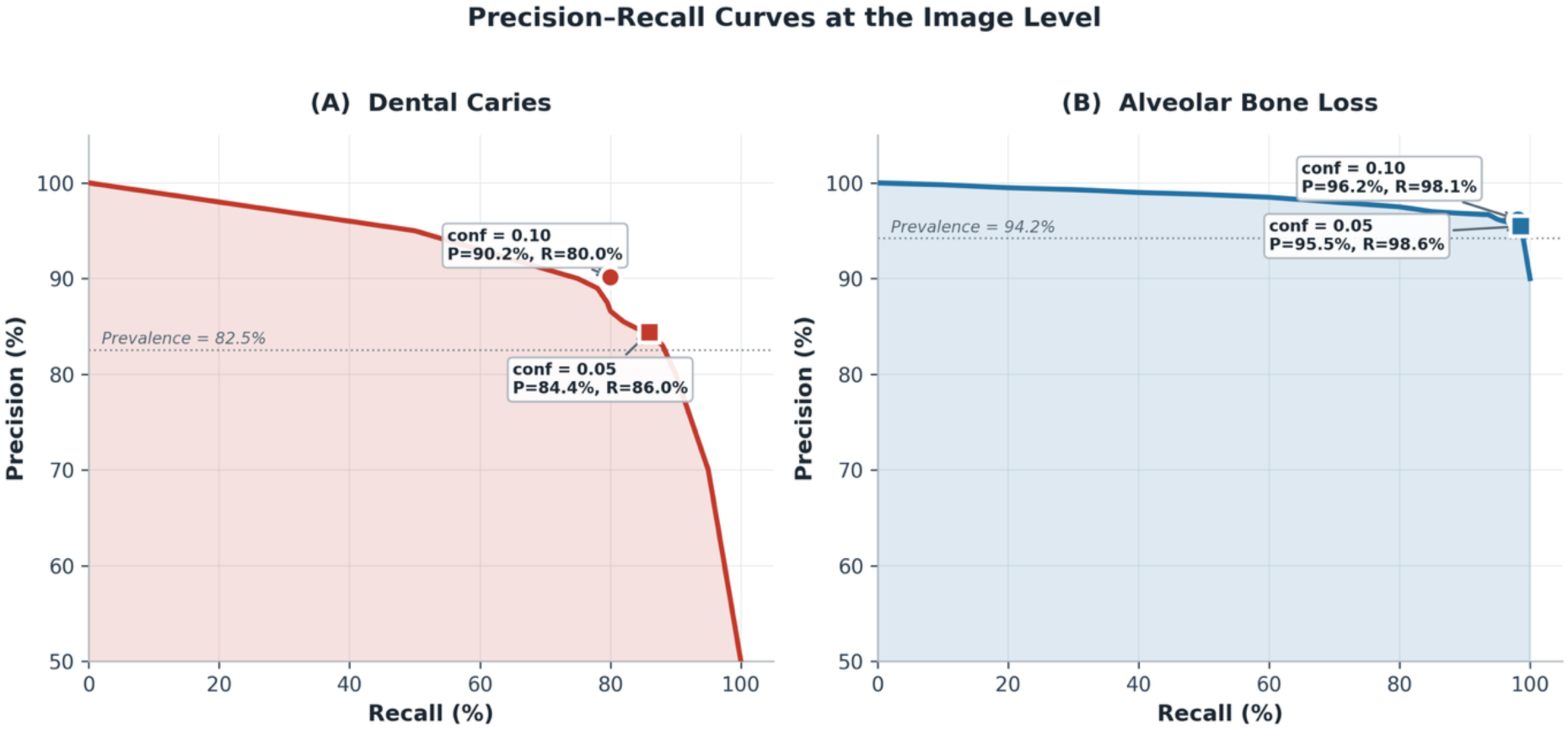
Precision-Recall curves for the caries and bone loss models. Precision-Recall curves for the caries (A) and bone loss (B) models at the image level. The curves show that both models, particularly the bone loss detector, *maintain high precision as recall increases, indicating robust performance at confidence thresholds of 0.10 and 0.05*.

## 4. Discussion

This study demonstrates that a YOLO based deep learning system can reliably detect both dental caries and alveolar bone loss on periapical and bitewing radiographs with high sensitivity. Our work is distinguished by three key methodological strengths. First, we developed and validated two dedicated detection models within a unified framework, enabling simultaneous screening for the two most prevalent oral diseases encountered in routine clinical practice. Second, we employed a rigorous 5-fold cross-validation strategy that confirmed the stability and reproducibility of our results across diverse data partitions, addressing a common limitation in the field where models are evaluated on a single, potentially favorable split. Third, we benchmarked our methodology and performance against the newly established ANSI/ADA Standard for dental AI validation, a step that, to our knowledge, has not been undertaken in any prior study. Both models not only met but substantially exceeded the Standard’s minimum quality threshold, providing an objective, standards-based confirmation of clinical readiness. Taken together, these results position the system as a viable ‘second reader’ capable of augmenting clinician decision-making during routine radiographic interpretation, reducing the risk of missed pathology in high-volume clinical settings

When compared to the existing literature (Table 3), our models demonstrated highly competitive performance. For caries detection, our recall of 86.0% is notably higher than both the pooled YOLO sensitivity of 79.3% reported in the Lam et al. (2025) meta-analysis [20] and the 79.8% recall of the comparable YOLOv8 study by Bayati et al. (2025) [12], while maintaining a comparable precision of 84.4%. Relative to earlier CNN-based approaches, our model had numerically higher performance metrics compared to Lee et al. (2021) [22], whose model achieved only 65.0% recall and 63.3% precision on a smaller dataset of 304 bitewing radiographs. Our accuracy of 83.0% is within the range of the landmark study by Lee et al. (2018) [8], which reported 82–89% accuracy using 3,000 periapical images with a GoogLeNet Inception v3 architecture. Given that our dataset is approximately one-third the size of that study, this comparable performance is encouraging and suggests that expanding the dataset through multi-site collaboration could yield further improvements.

**Table 3.** Comparison of performance metrics with recent deep learning studies for caries and bone loss detection.

| **Study** | **Year** | **Target** | **Model** | **Dataset** | **Key Performance Metric(s)** | **Our Model Comparison** |
| --- | --- | --- | --- | --- | --- | --- |
| *Our Study* | 2026 | Caries | YOLOv8s | 1,063 periapical radiographs | Recall: 86.0%, Precision: 84.4% | Baseline |
| *Lam et al. [20]* | 2025 | Caries | YOLO systematic review and meta-analysis | 15 studies | Pooled sensitivity: 79.3% | Our recall was 6.7 percentage points higher. |
| *Bayati et al. [12]* | 2025 | Caries | YOLOv8 | 552 bitewing radiographs | Recall: 79.8%, Precision: 84.8% | Our recall was 6.2 percentage points higher, while precision was 0.4 percentage points lower. |
| *Lee et al. [8]* | 2018 | Caries | GoogLeNet Inception-v3 | 3,000 periapical radiographs | Accuracy: 82.0%, AUC: 0.845 | Dataset and summary metrics verified; direct recall/precision comparison not available from the cited summary. |
| *Lee et al. [22]* | 2021 | Caries | U-Net | 304 bitewing radiographs | Recall: 65.0%, Precision: 63.3% | Our recall was 21.0 percentage points higher and precision was 21.1 percentage points higher. |
| *Bayırlı et al. [23]* | 2026 | Caries | YOLOv8x-seg | 1,197 bitewing radiographs | Precision: 65%, Recall: 50% | Our recall was 36.0 percentage points higher and precision was 19.4 percentage points higher. |
| *Mutlu et al. [25]* | 2026 | Caries | YOLOv11 and YOLOv12 | 2,297 panoramic radiographs | Recall: 83.7%, Precision: 85.4%, F1: 0.845 (internal test set, YOLOv11) | Our recall was 2.3 percentage points higher, while precision was 1.0 percentage point lower. |
| *Our Study* | 2026 | Bone Loss | YOLOv8s | 1,063 periapical radiographs | Recall: 98.6%, Precision: 95.5% | Baseline |
| *AlGhaihab et al. [21]* | 2025 | Bone Loss | Denti.AI | 22 periapical radiographs (123 tooth surfaces) | Sensitivity: 76% | Our recall was 22.6 percentage points higher. |
| *Krois et al. [10]* | 2019 | Bone Loss | CNN | 2,001 panoramic radiographs | Sensitivity: 81%, Specificity: 81% | Our recall was 17.6 percentage points higher. |
| *Bayırlı et al. [23]* | 2026 | Bone Loss | YOLOv8x-seg | 1,197 bitewing radiographs | Precision: 84%, Recall: 93% | Our recall was 5.6 percentage points higher and precision was 11.5 percentage points higher. |
| *Mutlu et al. [25]* | 2026 | Bone Loss | YOLOv11 and YOLOv12 | 2,297 panoramic radiographs | Recall: 71.0%, Precision: 69.6%, F1: 0.703 (internal test set, YOLOv11) | Our recall was 27.6 percentage points higher and precision was 25.9 percentage points higher. |

For bone loss detection, the contrast with the existing literature is even more pronounced. Our model’s recall of 98.6% substantially exceeds the 76% sensitivity reported by AlGhaihab et al. (2025) [21] using the commercial Denti.AI system on periapical radiographs, and the 81% sensitivity achieved by the seminal study of Krois et al. (2019) [10] on panoramic radiographs. It is important to acknowledge, however, that direct comparison across studies is inherently limited by differences in datasets, radiograph types (periapical vs. bitewing vs. panoramic), annotation protocols, and evaluation criteria. In particular, our use of a lenient IoU > 0 threshold, while clinically justified for a screening application, may contribute to higher recall values compared to studies employing the stricter mAP@0.5 standard.

The selection of a low confidence threshold (0.05) in this study was a deliberate design choice aligned with established practices for medical AI screening tools. As noted in recent literature, AI systems deployed for clinical triage are typically optimized for higher sensitivity and lower specificity compared to human readers, as the clinical cost of a false negative (missed pathology) significantly outweighs the cost of a false positive (which is subsequently reviewed by the clinician) [26]. Similarly, in dental AI, confidence thresholds as low as 0.001 have been employed in YOLO-based caries detection studies to maximize recall and minimize missed lesions [27]. By reporting performance across multiple thresholds (0.05 and 0.10) and providing full Precision–Recall curves, this study transparently illustrates this necessary clinical trade-off.

It is important to acknowledge, however, that optimizing for high sensitivity inherently carries a trade-off with specificity. La Rosa [29] highlighted that AI systems with high sensitivity but lower specificity are susceptible to false-positive results, raising concerns about potential overtreatment, particularly in populations where most early lesions do not progress. This concern reinforces the augmented intelligence paradigm adopted in this study: the AI system is not intended to trigger autonomous treatment decisions, but rather to flag areas of concern for subsequent clinician verification. In this workflow, a false positive is reviewed and dismissed by the dentist at minimal cost, whereas a false negative represents a missed lesion with potentially significant clinical consequences.

As AI technologies rapidly evolve, the critical need for standardized evaluation criteria has become paramount. The recent publication of the ANSI/ADA Standard No. 1110-1 provides a vital framework for validating AI image analysis systems, ensuring they meet rigorous ethical, technical, and quality assurance benchmarks before clinical adoption. This study not only demonstrates the high performance of a YOLOv8-based deep learning model but also rigorously aligns its methodology with this new ADA standard, providing a robust template for future dental AI research. To explicitly demonstrate this alignment, Table 1 provides a comprehensive cross-reference of our study’s methodology against the Quality Assurance criteria mandated by the ADA standard.

While this study achieves full compliance across the majority of the ANSI/ADA Standard criteria, two domains were designated as partially compliant, reflecting the pragmatic constraints of current feasibility research. Regarding Domain 2 (Machine Translation), the standard’s novel requirement for ‘AI Hallucination Detection’ was addressed through rigorous false-positive analysis via Precision metrics. However, it is important to emphasize that this study represents a proof-of-concept; there are currently no plans to deploy this algorithm in a clinical setting. Before deployment can be considered, the dataset must be expanded, additional clinical conditions must be incorporated, and explicit hallucination monitoring protocols must be developed for the real-world deployment phase. Critically, the envisioned deployment model is one of augmented intelligence, not autonomous diagnosis. The system is designed to function as a clinical decision support tool in which the dentist always cross-checks every AI detection. Under this paradigm, a false positive carries minimal clinical risk, it simply prompts the clinician to examine a region that is ultimately healthy, adding only a brief moment of verification. In contrast, a false negative (a missed lesion) carries significant clinical risk, as it may lead to delayed treatment. Therefore, the system’s optimization for high sensitivity is deliberately aligned with this augmented workflow: it is far better for the AI to flag a questionable area for the dentist to verify than to miss a true lesion entirely. Regarding Domain 4 (Data Collection Quality), formal Inter-Annotator Agreement (IAA) statistics were not calculated. Instead, a highly robust cross-over consensus methodology was utilized. Two experienced dentists independently annotated the entire dataset, exchanged and reviewed each other’s work, and then jointly double-checked all labels, discussing any discrepancies until full consensus was reached. This rigorous approach ensures a highly reliable ground truth that aligns with the spirit of the ADA’s data quality requirements.

The clinical implication of such a tool is a streamlined and more reliable diagnostic workflow. The AI can act as a tireless ‘second opinion,’ highlighting potential areas of concern that a clinician might have overlooked due to fatigue or the subtlety of the finding. This can lead to earlier treatment, better patient outcomes, and a reduction in the variability of care. As La Rosa [29] emphasized, the role of clinicians remains essential in assessing patient risk and making informed therapeutic decisions; new AI-based systems should be seen as tools to support the clinician’s expertise in this decision-making process, not to replace it. This philosophy is foundational to our system design.

Despite the high performance of the YOLOv8 models demonstrated in this study, the integration of AI into clinical workflows warrants cautious optimism. Recent literature has raised alarming concerns regarding ‘automation bias’ and ‘confidence inflation’ in medical AI. A 2026 study in npj Digital Medicine demonstrated that when novice medical learners were exposed to misleading AI explanations, their diagnostic accuracy significantly degraded, yet their confidence in those incorrect diagnoses paradoxically increased [28]. This phenomenon, where clinicians commit errors with AI-induced conviction, highlights the danger of deploying AI as an unquestioned oracle. It reinforces the necessity of the ‘augmented intelligence’ paradigm advocated in this study, where the AI serves strictly as a ‘second reader’ to flag potential areas of concern, rather than providing definitive diagnostic explanations that might unduly steer clinical reasoning. Furthermore, it underscores the importance of our educational initiative (Section 1); as AI tools become ubiquitous, dental curricula must evolve beyond teaching students how to use AI, to explicitly training them in AI-auditing, critical appraisal, and skepticism to prevent high-confidence commission errors.

This study should be viewed as a successful feasibility study conducted within a single academic institution. The primary limitation is the use of a single-site dataset, which may not fully represent the diversity of imaging equipment and patient populations found in a multi-center study. Furthermore, while our dataset of 1,063 images is a moderate sample size, deep learning models invariably benefit from larger and more diverse datasets. We anticipate that the performance of the caries detection model, in particular, would improve with a larger sample size, as caries lesions can present with more subtle and varied appearances than bone loss. Additionally, in the context of the ANSI/ADA Standard, two specific limitations must be acknowledged. First, while annotations were performed by two experienced practitioners, formal Inter-Annotator Agreement (IAA) metrics were not calculated; instead, discrepancies were resolved via consensus. Future studies must quantify IAA to fully satisfy the standard’s data labeling quality requirements. Second, the reliance on EHR screenshots rather than native DICOM files limits the referential integrity and provenance tracking mandated by the standard. Future multi-site collaborations will prioritize the collection of native DICOM images to ensure full compliance with the ADA’s image annotation pedigree requirements.

Future work will focus on addressing these limitations by expanding this project into a multi-site collaboration. By incorporating data from other institutions, we can build a more generalized model that is robust to variations in imaging protocols and patient demographics. This will be a critical step in validating the real-world clinical utility of the system and moving towards regulatory approval and broader clinical adoption.

## 5. Conclusion

This study demonstrates that a YOLO based deep learning system can support the image-level screening of dental caries and alveolar bone loss on periapical and bitewing radiographs with high sensitivity. The results of this feasibility study provide a strong foundation for future multi-site research. By integrating this type of AI development into the dental curriculum, this project also served as a valuable capacity-building exercise, preparing the next generation of dental professionals for a future in which AI is an integral part of clinical practice.

## Data Availability

Data Availability: The data that support the findings of this study are not publicly available due to patient privacy restrictions under the IRB-approved protocol. Requests for access to the de-identified dataset may be directed to the corresponding author.

## Acknowledgements

The authors wish to acknowledge the invaluable contributions of those who assisted with data collection, which played a critical role in this research. Special thanks are extended to Adonis Ponciano, Akshaya Raviraj, Ariana Margolis, Danielle Smolyar, Michelle Zak, and Zhyear Othman for their dedicated efforts.

